# Geographic and Temporal Patterns in Covid-19 Mortality by Race and Ethnicity in the United States from March 2020 to February 2022

**DOI:** 10.1101/2022.07.20.22277872

**Authors:** Dielle J. Lundberg, Ahyoung Cho, Rafeya Raquib, Elaine O. Nsoesie, Elizabeth Wrigley-Field, Andrew C. Stokes

**Affiliations:** Department of Global Health, Boston University School of Public Health; Center for Antiracist Research, Boston University; Department of Political Science, Boston University; Department of Sociology and Minnesota Population Center, University of Minnesota

**Keywords:** Covid-19 Mortality, Racial/Ethnic Disparities, Urban-Rural Differences, Covid-19 Vaccination, Structural Racism

## Abstract

Prior research has established that American Indian, Alaska Native, Black, Hispanic, and Pacific Islander populations in the United States have experienced substantially higher mortality rates from Covid-19 compared to non-Hispanic white residents during the first year of the pandemic. What remains less clear is how mortality rates have changed for each of these racial/ethnic groups during 2021, given the increasing prevalence of vaccination. In particular, it is unknown how these changes in mortality have varied geographically. In this study, we used provisional data from the National Center for Health Statistics (NCHS) to produce age-standardized estimates of Covid-19 mortality by race/ethnicity in the United States from March 2020 to February 2022 in each metro-nonmetro category, Census region, and Census division. We calculated changes in mortality rates between the first and second years of the pandemic and examined mortality changes by month. We found that when Covid-19 first affected a geographic area, non-Hispanic Black and Hispanic populations experienced extremely high levels of Covid-19 mortality and racial/ethnic inequity that were not repeated at any other time during the pandemic. Between the first and second year of the pandemic, racial/ethnic inequities in Covid-19 mortality decreased—but were not eliminated—for Hispanic, non-Hispanic Black, and non-Hispanic AIAN residents. These inequities decreased due to reductions in mortality for these populations alongside increases in non-Hispanic white mortality. Though racial/ethnic inequities in Covid-19 mortality decreased, substantial inequities still existed in most geographic areas during the pandemic’s second year: Non-Hispanic Black, non-Hispanic AIAN, and Hispanic residents reported higher Covid-19 death rates in rural areas than in urban areas, indicating that these communities are facing serious public health challenges. At the same time, the non-Hispanic white mortality rate worsened in rural areas during the second year of the pandemic, suggesting there may be unique factors driving mortality in this population. Finally, vaccination rates were associated with reductions in Covid-19 mortality for Hispanic, non-Hispanic Black, and non-Hispanic white residents, and increased vaccination may have contributed to the decreases in racial/ethnic inequities in Covid-19 mortality observed during the second year of the pandemic. Despite reductions in mortality, Covid-19 mortality remained elevated in nonmetro areas and increased for some racial/ethnic groups, highlighting the need for increased vaccination delivery and equitable public health measures especially in rural communities. Taken together, these findings highlight the continued need to prioritize health equity in the pandemic response and to modify the structures and policies through which systemic racism operates and has generated racial health inequities.

## INTRODUCTION

The coronavirus disease (Covid-19) pandemic has resulted in substantial loss-of-life and exacerbated long-existing racial/ethnic inequities in mortality across the United States.^1–5^ While life expectancy declined by 1.5 years during 2020 in the overall population, some racial/ethnic groups experienced more substantial decreases in life expectancy, such as a decline of 3.0 years for the Hispanic population, a decline of 2.9 years for the non-Hispanic Black population, and a decline of 4.5 years for American Indian or Alaska Native (AIAN) populations.^6,7^

Considerable prior research has documented racial/ethnic inequities in mortality from Covid-19 during the pandemic.^8–15^ During 2020, age-standardized death rates from Covid-19 were 2.6 times higher for AIAN populations than for the white population.^16^ Other racial/ethnic groups also experienced higher death rates than the white population, including Hispanic residents (2.3 times higher), Black residents (2.1 times higher), and Native Hawaian or Other Pacific Islander (NHOPI) residents (1.7 times higher). Asian American residents reported similar age-standardized death rates to white residents.^16^

Structural racism – “the totality of ways in which societies foster racial discrimination, via mutually reinforcing inequitable systems… that in turn reinforce discriminatory beliefs, values, and distribution of resources”^17^ – is strongly associated with racial/ethnic inequities in Covid-19 mortality, suggesting that structural racism has driven differences in mortality during the pandemic.^18–20^ The lasting impacts of chattel slavery, Jim Crow era policies, the genocide of Indigenous peoples, the occupation of Mexican territories, and other state-sanctioned violence have led to contemporary fundamental causes of health such as residential segregation, occupational segregation, policing, incarceration, health care access, and medical racism.^17,21^ Prior literature has demonstrated that each of these systems is responsible for increasing racial/ethnic inequities in health and mortality in the years before the pandemic.^22–28^

During the pandemic, multiple mechanisms have been identified through which structural racism has further increased racial/ethnic inequities in mortality, including differential infection rates, weathering, and experiences in health care.^29–31^ First, increased Covid-19 infection rates for Black, Hispanic, AIAN, and NHOPI populations may relate to multiple factors, such as occupational segregation.^32^ From the start of the pandemic, white workers were more likely to be able to work from home compared to Black and Hispanic workers and were under-represented in professions with high risk of exposure to Covid-19.^33^ Household crowding, which relates both to residential segregation and to cultural differences around multigenerational households, was also greater for Black and Hispanic populations and was associated with Covid-19 infection.^9,33,34^ Second, weathering refers to the process of lifelong exposure to structural racism and other systemic stressors, which over time lead to poorer health and increased risk of underlying health conditions such as diabetes, cardiovascular disease and obesity.^29,35^ As a result of weathering, Black, Hispanic, AIAN, Asian, and NHOPI populations report higher rates of underlying health conditions that increase risk for Covid-19 mortality.^36–38^ Finally, significant racial/ethnic inequities exist in access to health insurance and health care.^28,39^ Disparities in Black/white mortality from Covid-19 have been shown to relate to hospital quality.^40^ White individuals were more likely to access higher-quality hospitals during the pandemic, whereas Black individuals were more likely to access lower-quality hospitals, due to hospital and residential segregation.^41–43^ These hospitals are believed to provide lower-quality care because of differences in payer mix and systemic under-funding.^40^

While racial/ethnic inequities in Covid-19 mortality have been extensively studied during the first year of the pandemic, less is known about how mortality patterns have changed from 2020 to 2021, given the increasing prevalence of vaccination and the changing geography of the pandemic. At the national level, racial/ethnic disparities decreased substantially between 2020 and 2021; however, this was not only because Black, Hispanic, and Asian death rates decreased but also because white death rates increased from 66 deaths per 100,000 residents in 2020 to 90 deaths per 100,000 residents in 2021.^16^

There are several reasons to anticipate that the racial/ethnic patterning of Covid-19 mortality may have changed in tandem with changing geographic risk patterns during the second year of the pandemic. First, the pandemic moved to increasingly rural areas in 2021. In the Far West, Great Lakes, Mideast, and New England regions, mortality rates were higher in urban areas in 2020 but higher in rural areas in 2021. In the Southeast, Southwest, Rocky Mountain, and Plains regions, mortality rates were higher in rural areas in 2020 but grew even higher in 2021. Second, preliminary evidence has suggested that Covid-19 mortality rates have been elevated among Black residents in nonmetro areas, indicating that rural areas may be an area of increased concern for Covid-19 mortality among Black residents and other racial/ethnic groups that have been disproportionately affected by the pandemic.^8^ Third, differences in vaccine uptake have been reported by race/ethnicity and metro-nonmetro category. While some Black adults had mistrust of the Covid-19 vaccine initially due in part to a history of medical racism, vaccine hesitancy decreased more quickly among Black adults once vaccination efforts began.^44^ Furthermore, a poll in mid-2021 showed that the most vaccine hesitant group was older, white individuals who identified as Republican.^45^ Vaccine uptake was also lower among rural than urban residents in the first half of 2021, suggesting that mortality may have worsened among residents in rural areas.^46^ Finally, many AIAN, Black, Hispanic, Asian American, and NHOPI communities have responded in highly coordinated ways to the pandemic, with community-led campaigns for vaccination and other protective efforts.^47^ These communities may have experienced reductions in mortality as the pandemic progressed.

In this study, we produced age-standardized estimates of Covid-19 mortality by race/ethnicity in the United States from March 2020 to February 2022 in each metro-nonmetro category, Census region, and Census Division. These estimates allowed us to examine changes in mortality rates between the first and second years of the pandemic and to explore patterns of mortality by month. Such an analysis provides an opportunity to identify geographic areas where mortality has increased and decreased during the pandemic. Finally, we assessed how vaccination rates were associated with changes in mortality rates by race/ethnicity to explore the role vaccination has had in changing Covid-19 death rates across the country.

## METHODS

### Data

#### Mortality Data

We used multiple cause of death data from the National Center for Health Statistics (NCHS) obtained via the Centers for Disease Control and Prevention (CDC) WONDER online database. Data from 2020 were final, whereas data from 2021 and 2022 were provisional and may be incomplete and subject to revision. Data was last updated on July 6, 2022. We used ICD-10 code U07.1 to identify deaths where Covid-19 was listed anywhere on the death certificate.

Race/ethnicity was presented in single race categories (Hispanic, non-Hispanic AIAN, non-Hispanic Asian, non-Hispanic NHOPI, non-Hispanic Black, non-Hispanic multi-racial, and non-Hispanic white). Hereafter, we omit the non-Hispanic label (e.g., “Black” for “non-Hispanic Black”) for parsimony. We excluded the non-Hispanic multi-racial category from the analysis because it was a heterogeneous group and would reflect distinct populations in different geographic locations.

Year one of the pandemic consisted of the months from March 2020 to February 2021. Year two of the pandemic consisted of the months from March 2021 to February 2022.

Age was provided in 10-year age intervals. Since CDC WONDER suppresses death counts between 1 and 9, we condensed the 10-year age intervals into 3 age groups (0-54, 55-74, and 75+) to eliminate data suppression that would have otherwise occurred, especially in racial/ethnic groups with smaller populations. We also condensed the 2013 Urbanization categories into 3 metro-nonmetro categories of residence – large metro (large centro metro and large fringe metro), medium/small metro (medium metro and small metro), and nonmetro (micropolitan (nonmetro) and noncore (nonmetro)) – to avoid data suppression.

We queried Covid-19 mortality by race/ethnicity and pandemic year across 3 metro-nonmetro categories and 4 Census regions. For the 3 racial/ethnic groups that were populous enough to avoid data suppression (non-Hispanic Black. Hispanic and non-Hispanic white), we queried Covid-19 mortality by race/ethnicity and month across the 3 metro-nonmetro categories and 4 Census regions. We also queried Covid-19 mortality by race/ethnicity, year and combinations of metro-nonmetro category and Census region along with combinations of metro-nonmetro category and Census division. Our detailed approach to querying data from CDC WONDER to minimize data suppression is provided in the **Methods Supplement**.

#### Population Data

We used mid-year 2020 and 2021 population estimates from the U.S. Census Bureau. These included estimates for 6 racial/ethnic groups (Hispanic, non-Hispanic AIAN alone, non-Hispanic Asian alone, non-Hispanic NHOPI alone, non-Hispanic Black alone, and non-Hispanic white alone) and 3 age groups (0-54, 55-74, and 75+). We aggregated these estimates from the county-level to the levels of metro-nonmetro categories, Census regions, and Census divisions. Population estimates for 2022 were not available, so population estimates for 2021 were used for January and February 2022 in the analysis.

#### Vaccination Data

We used CDC data on the percent of the population fully vaccinated (defined as having 2 doses of Pfizer-BioNTech, 2 doses of Moderna, or 1 dose of Johnson & Johnson) in a county, as of February 28, 2022. Data on vaccination rates by race/ethnicity were not available at the county-level or in all states, so we examined vaccination rates in the overall population. We aggregated the county-level data on vaccination in the overall population to combinations of metro-nonmetro categories and Census divisions.

### Data Imputation

While our data extraction procedure was designed to minimize data suppression, a small number of cells, especially for less populous racial/ethnic groups and nonmetro areas, were imputed because values between 1 and 9 were suppressed. We imputed missing values with random draws from a beta distribution (1,1) and multiplied the results by 10 to satisfy the range of death counts (1 to 9). Less than 5% of data was imputed throughout the analysis, and data that were imputed are indicated in each table and figure.

### Age-Standardized Death Rates

Since racial/ethnic groups in the United States have different age distributions, age standardization is necessary to produce accurate comparisons of mortality by race/ethnicity and estimates of racial/ethnic disparities.^11,20,48^ To produce age-standardized death rates, we first generated age-specific death rates for each racial/ethnic group at each geographic and temporal unit for three age groups (0-54, 55-74, and 75+). We then calculated the percentage of the 2020 U.S. population that belonged to each of these age groups. We multiplied each age-specific death rate by the corresponding percentage of the U.S. population in that age group and then summed the 3 components to produce the age-standardized death rate. Further details about the age-standardization procedure are provided in the **Methods Supplement**.

We calculated age-standardized death rates, changes in age-standardized death rates between the first and second year of the pandemic, rate ratios comparing non-Hispanic AIAN, non-Hispanic Asian, non-Hispanic Black, non-Hispanic NHOPI, and Hispanic residents to non-Hispanic white residents, and changes in rate ratios between the first and second year of the pandemic. We assessed these patterns across several geographic units (metro-nonmetro categories, Census regions, and Census divisions) and temporal units (years and months). We also explored differences during different waves of the pandemic – the early pandemic (March to August 2020), alpha wave (September 2020 to May 2021), delta wave (June to October 2021), and omicron wave (November 2021 to February 2022). For the more granular geographic or temporal units, we limited to the three largest racial/ethnic groups.

### Analysis

Next, we examined the relationship between the percent fully vaccinated in an area and changes in age-standardized mortality rates between the first and second year of the pandemic using a linear regression model. The unit for this analysis was combinations of metro-nonmetro categories and Census divisions. We generated three models for Hispanic, non-Hispanic Black, and non-Hispanic white residents. The regressions were weighted by the population of the specific racial/ethnic group for each geographic unit in 2020.

This study used de-identified publicly available data and was exempted from review by the Boston University Medical Center Institutional Review Board. Programming code was developed using R, version 3.6.3, and replication code is available online on GitHub (https://github.com/BU-Center-for-Antiracist-Research/county-level-racial-ethnic-inequities-in-COVID-19-mortality).

## RESULTS

### Covid-19 Mortality across Metro-Nonmetro Categories

**Table 1** shows changes in age-standardized Covid-19 mortality rates across metro-nonmetro categories from the first year of the pandemic (March 2020 to February 2021) to the second year (March 2021 to February 2022). These patterns are also visualized in **Figure 1**. During the first year of the pandemic, Covid-19 death rates were higher in nonmetro areas for non-Hispanic Black, non-Hispanic AIAN, and non-Hispanic white residents than in large metros and medium/small metros. In contrast, Covid-19 death rates were higher in large metro areas for non-Hispanic Asian and non-Hispanic NHOPI than in nonmetro areas and medium/small metros. Hispanic death rates were relatively similar across metro-nonmetro categories.

**Table 1.**
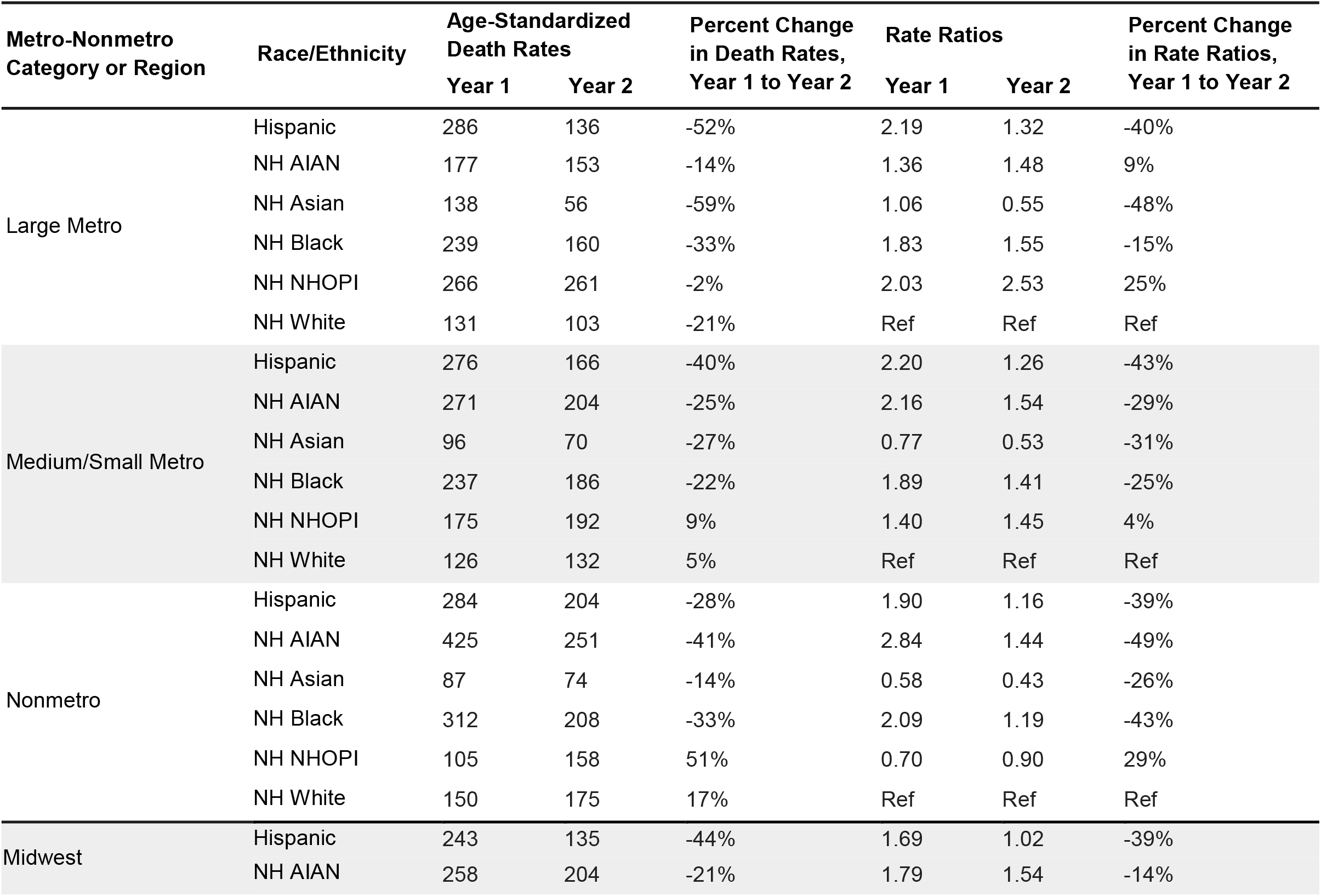
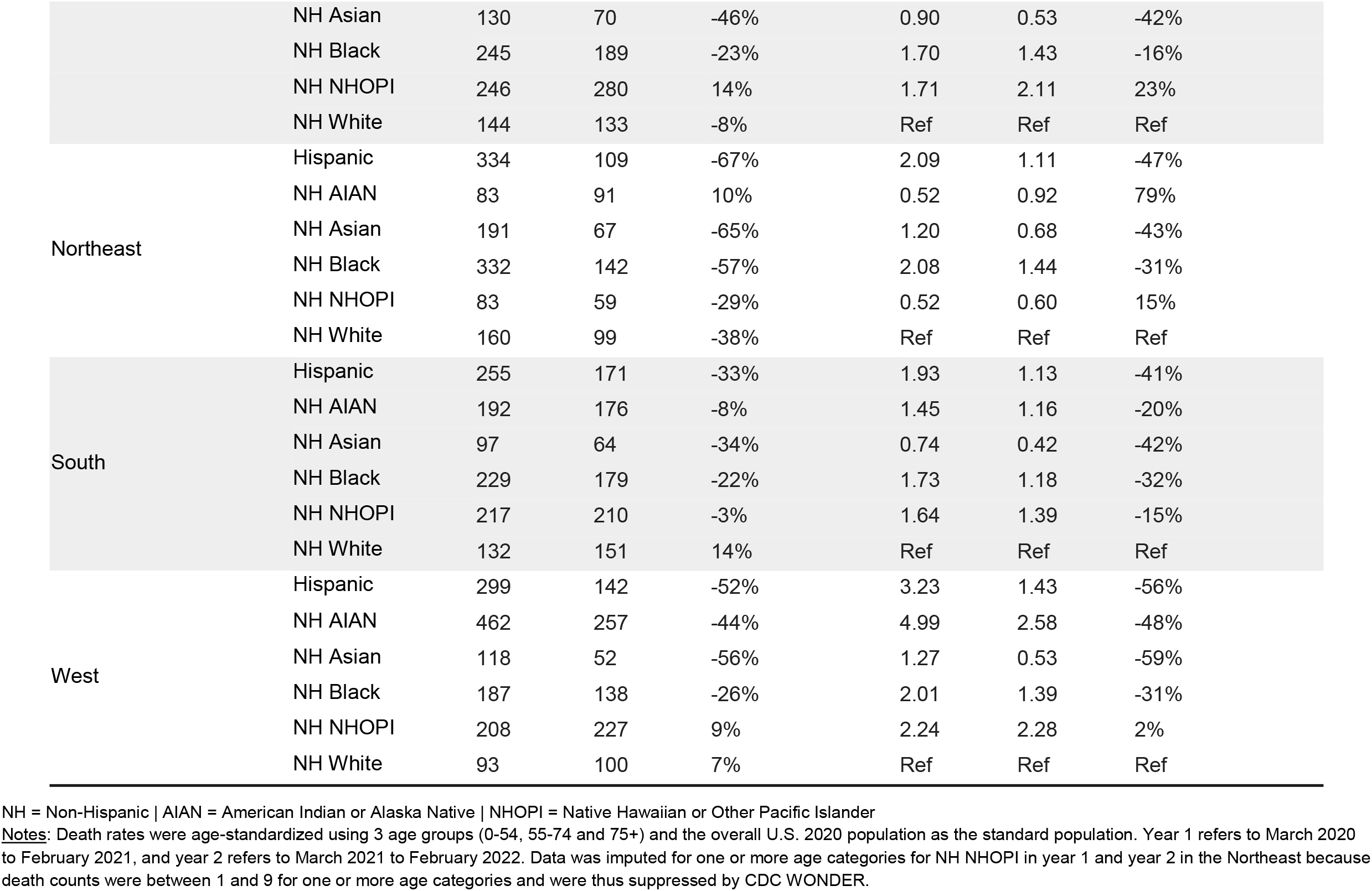
Changes in Age-Standardized Covid-19 Death Rates by Race/Ethnicity across Metro-Nonmetro Categories and Census Regions, March 2020-February 2021 to March 2021-February 2022

**Figure 1.**
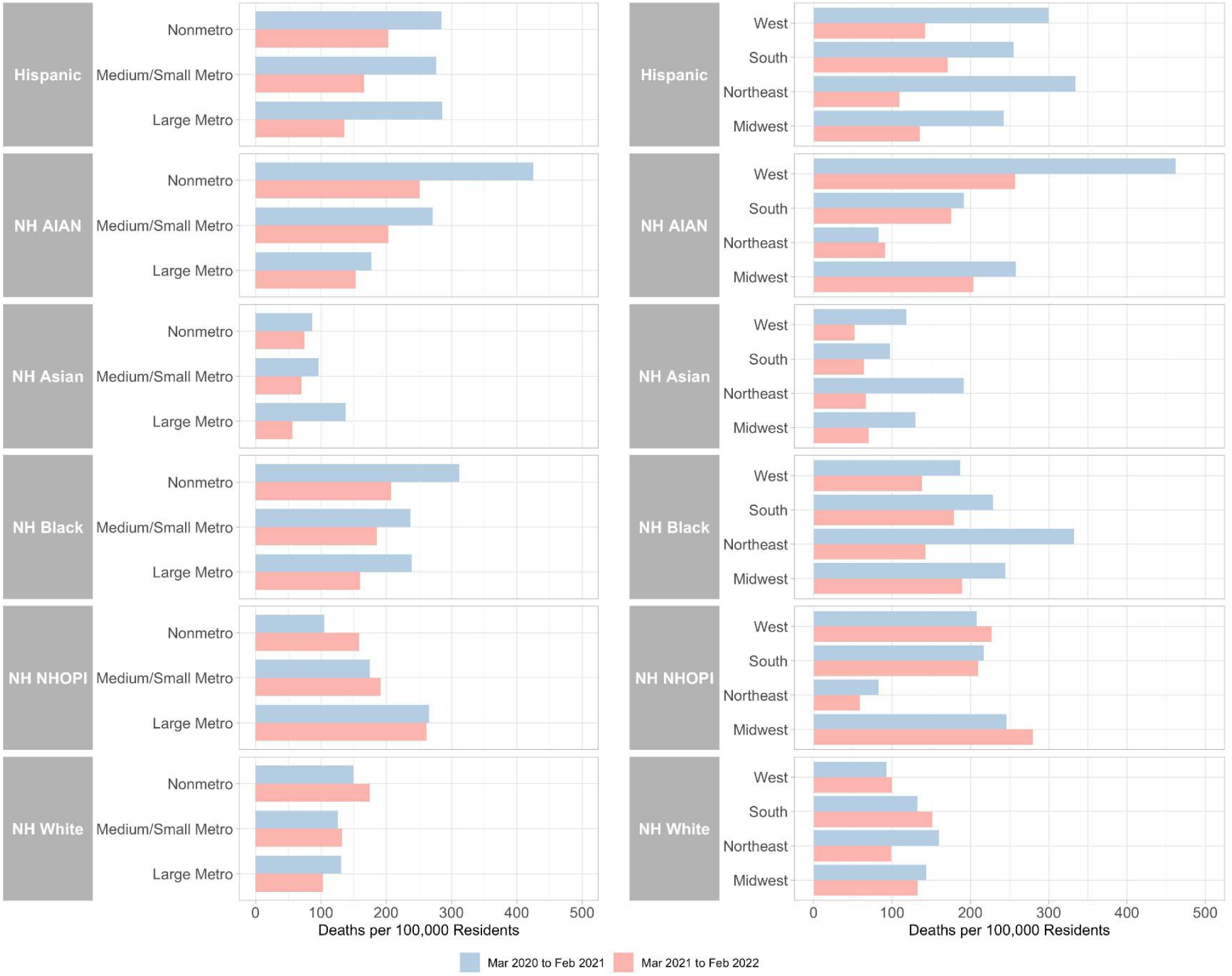
Age-Standardized Covid-19 Death Rates by Race/Ethnicity across Metro-Nonmetro Categories and Census Regions, March 2020-February 2021 and March 2021-February 2022 NH = Non-Hispanic | AIAN = American Indian or Alaska Native | NHOPI = Native Hawaiian or Other Pacific Islander <u>Notes:</u> Death rates were age-standardized using 3 age groups (0-54, 55-74 and 75+) and the overall U.S. 2020 population as the standard population. Data was imputed for one or more age categories for NH NHOPI in year 1 and year 2 in the Northeast because death counts were between 1 and 9 for one or more age categories and were thus suppressed by CDC WONDER.

During the first year of the pandemic, rate ratios for Hispanic residents compared to white residents were greater in medium/small metros (2.20) and large metros (2.19) than in nonmetros (1.90). Rate ratios for non-Hispanic Black residents were greater in nonmetro areas (2.09) than medium/small metro areas (1.89) and large metros (1.83). Rate ratios were the greatest among any racial/ethnic group for non-Hispanic AIAN residents in nonmetro areas (2.84). Non-Hispanic AIAN death rates varied substantially across metro-nonmetro categories, with rate ratios of 2.16 in medium/small metros and 1.36 in large metros. Non-Hispanic Asian death rates were lower than non-Hispanic white death rates in nonmetro areas and medium/small metro areas but not in large metro areas.

Between the first and second year of the pandemic, mortality rates decreased for non-Hispanic Black, non-Hispanic AIAN, Hispanic, and non-Hispanic Asian populations across all metro-nonmetro categories. The relative decreases were greatest for non-Hispanic Asian residents (-59%) in large metros and were also very large for non-Hispanic AIAN residents (-41%) in nonmetro areas. Mortality rates decreased more for Hispanic residents in large metros (-52%) than for Hispanic residents in medium/small metros (-40%) and nonmetro areas (-28%). Non-Hispanic white mortality rates increased but only in medium/small metros (5%) and nonmetro areas (17%). Similarly, mortality rates increased for non-Hispanic NHOPI residents but only in medium/small metros (9%) and nonmetro areas (51%).

Despite the noted decrease in mortality during the second year of the pandemic, Hispanic, non-Hispanic Black, non-Hispanic AIAN and non-Hispanic NHOPI residents in large metros had higher mortality rates than non-Hispanic white residents. In nonmetro areas, only Hispanic, non-Hispanic Black, and non-Hispanic AIAN residents had higher mortality rates than white residents. Rate ratios were generally smaller in the second year of the pandemic than the first.

### Covid-19 Mortality across Regions

**Table 2** shows changes in age-standardized Covid-19 mortality rates across Census regions from the first year of the pandemic to the second year. These patterns are also visualized in **Figure 1**. During the first year of the pandemic, Hispanic mortality rates were the highest in the Northeast followed by the West. Non-Hispanic Black and non-Hispanic white mortality rates were the highest in the Northeast followed by the Midwest and the South. Non-Hispanic Asian mortality rates were highest in the Northeast followed by the Midwest and the West. The Non-Hispanic AIAN mortality rate was the highest of any racial/ethnic group or region in the West, and the non-Hispanic NHOPI mortality rate was highest in the South followed by the West.

During the first year of the pandemic, rate ratios for Hispanic residents compared to non-Hispanic white residents were highest in the West (3.23) followed by the Northeast (2.09). Rate ratios for non-Hispanic Black residents were highest in the Northeast (2.08) followed by the West (2.01). Rate ratios for non-Hispanic AIAN residents were highest in the West (4.99) and Midwest (1.79). Non-Hispanic Asian mortality rates were higher than non-Hispanic white mortality rates in the West (1.27) and the Northeast (1.20). The rate ratio for non-Hispanic NHOPI residents was highest in the West (2.24).

Between the first and second year of the pandemic, mortality rates decreased for Hispanic, non-Hispanic Black, and non-Hispanic Asian residents across all regions. Non-Hispanic AIAN mortality rates decreased in the West (-44%), the Midwest (-21%), and the South (-8%) but increased in the Northeast (10%). Non-Hispanic white mortality rates increased in the South (14%) and West (7%) but decreased in the Northeast (-38%) and Midwest (-8%). Non-Hispanic NHOPI mortality rates also increased in the Midwest (14%) and West (9%).

During the second year of the pandemic, Hispanic and non-Hispanic Black residents across all regions had higher mortality rates than non-Hispanic white residents. Non-Hispanic AIAN residents had higher mortality rates than non-Hispanic white residents in the West (2.58), the Midwest (1.54), and the South (1.16). Non-Hispanic Asian residents had lower mortality rates than non-Hispanic white residents across all regions.

### Covid-19 Mortality across Combinations of Metro-Nonmetro Categories and Regions

Figure 2 and **Table S1** show changes in age-standardized Covid-19 mortality rates for Hispanic, non-Hispanic Black, and non-Hispanic white residents across combinations of metro-nonmetro categories and Census regions from the first year of the pandemic to the second year. During the first year, Hispanic, non-Hispanic Black, and non-Hispanic white mortality rates were all highest in large metros in the Northeast or nonmetro areas in the South.

**Figure 2.**
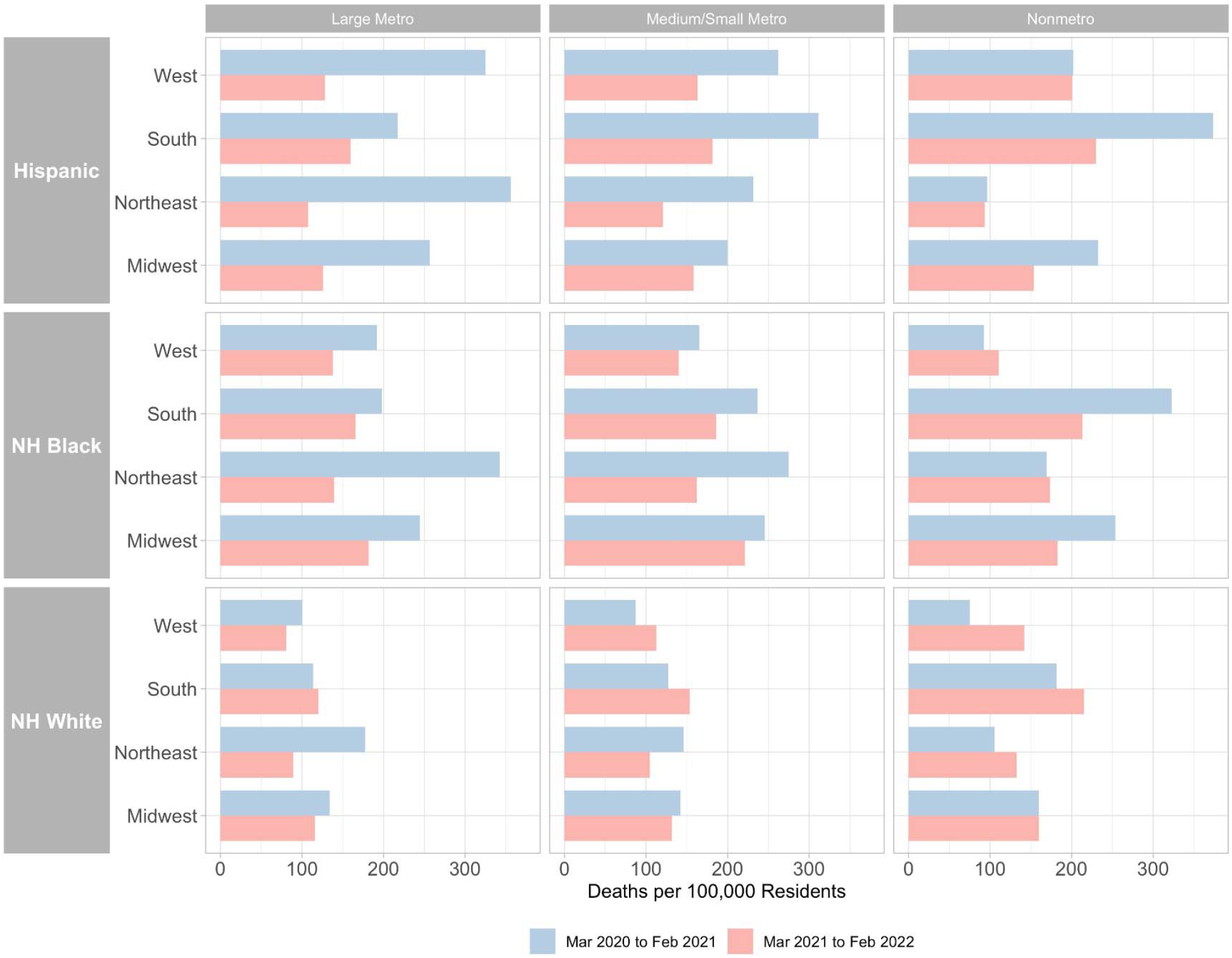
Age-Standardized Covid-19 Death Rates by Race/Ethnicity across Combinations of Census Regions and Metro-Nonmetro Categories, March 2020-February 2021 and March 2021-February 2022 NH = Non-Hispanic <u>Notes:</u> Death rates were age-standardized using 3 age groups (0-54, 55-74 and 75+) and the overall U.S. 2020 population as the standard population. Non-Hispanic NHOPI, Asian, and AIAN were excluded from this figure due to significant data suppression at this level of geographic detail. Data was imputed for one or more age categories for NH Black in year 1 in nonmetro areas in the Northeast and West and for Hispanic in year 1 in nonmetro areas in the Northeast because death counts were between 1 and 9 for one or more age categories and were thus suppressed by CDC WONDER.

During the first year of the pandemic, rate ratios were highest for Hispanic residents compared to non-Hispanic white residents in large metros (3.23), medium/small metros (3.01), and nonmetro areas (2.69) in the West. Rate ratios were highest for non-Hispanic Black residents in large metros (1.93) and medium/small metros (1.88) in the Northeast.

Between the first and second year of the pandemic, mortality rates decreased for Hispanic and non-Hispanic Black residents in all combinations of metro-nonmetro categories and regions except for non-Hispanic Black residents in nonmetro areas in the West (19%) and non-Hispanic Black residents in nonmetro areas in the Northeast (3%) where mortality rates increased. Non-Hispanic white mortality rates also decreased in most combinations of metro-nonmetro categories and regions except for nonmetro areas in the West (89%), medium/small metros in the West (29%), nonmetro areas in the Northeast (26%), medium/small metros in the South (21%), nonmetro areas in the South (18%), and large metros in the South (6%) where mortality rates increased.

Figure 3 shows changes in age-standardized mortality rates between the first and second year of the pandemic among combinations of metro-nonmetro categories and Census divisions. Hispanic mortality rates decreased the most in large metros in the Middle Atlantic, New England, and Pacific divisions. Non-Hispanic Black mortality rates decreased the most in New England and Middle Atlantic large metros followed by New England medium/small metros. Non-Hispanic white mortality rates increased the most in Pacific, Mountain, South Atlantic, and East South Central nonmetro areas. Mortality rates generally increased or decreased for all 3 racial/ethnic groups concurrently, but there were some geographic areas where mortality rates increased for non-Hispanic white residents and decreased for Hispanic and/or non-Hispanic Black residents such as Pacific medium/small metros, South Atlantic nonmetro areas, and South Atlantic medium/small metros.

**Figure 3.**
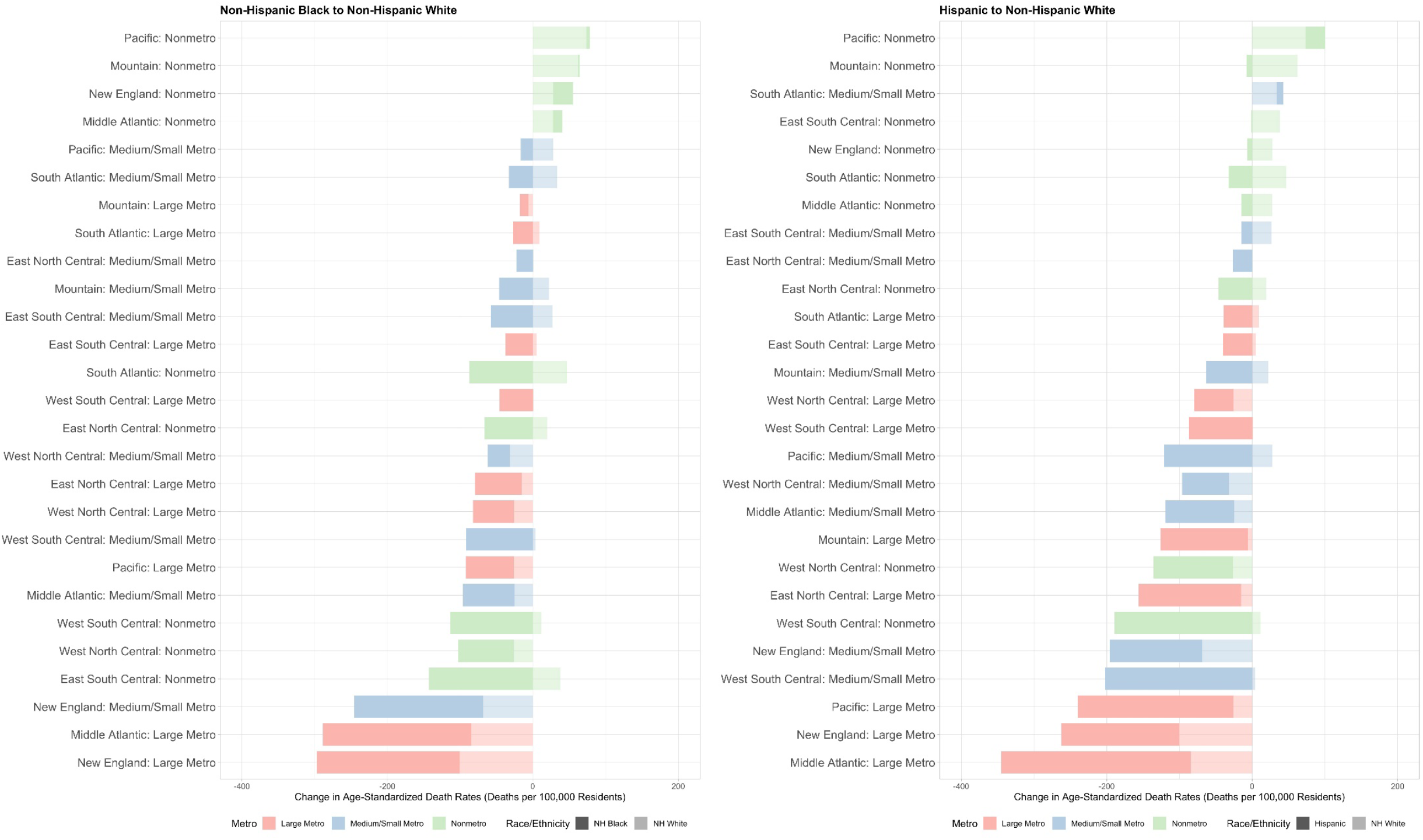
Changes in Age-Standardized Covid-19 Death Rates by Race/Ethnicity across Combinations of Census Divisions and Metro-Nonmetro Categories, March 2020-February 2021 to March 2021-February 2022 NH = Non-Hispanic <u>Notes:</u> Death rates were age-standardized using 3 age groups (0-54, 55-74 and 75+) using the overall U.S. 2020 population as the standard population. The unit in this analysis is combinations of metro-nonmetro categories and Census divisions (i.e. large metro Middle Atlantic). Data was imputed for one or more age categories for Hispanic residents in year 1 and year 2 in nonmetro areas in New England, Hispanic residents in year 1 in nonmetro areas in the Middle Atlantic division, NH Black residents in year 1 in nonmetro areas in the Middle Atlantic division, and NH Black residents in year 1 and year 2 in nonmetro areas in the Mountain division, New England, and the Pacific division because death counts were between 1 and 9 for one or more age categories and were thus suppressed by CDC WONDER.

During the second year of the pandemic, rate ratios were highest for Hispanic residents compared to non-Hispanic white residents in large metros (1.59), medium/small metros (1.45), and nonmetro areas (1.41) in the West. Rate ratios were highest for non-Hispanic Black residents in large metros in the West (1.71), medium/small metros in the Midwest (1.68), large metros in the Northeast (1.57), and medium/small metros in the Northeast (1.55).

### Covid-19 Mortality across Pandemic Waves

Figure 4 shows age-standardized Covid-19 mortality rates by month for Hispanic, non-Hispanic Black, and non-Hispanic white residents during the early pandemic (March to August 2020), alpha wave (September 2020 to May 2021), delta wave (June to October 2021), and omicron wave (November 2021 to February 2022) across metro-nonmetro categories. **Figure S1** shows the same patterns by Census region.

**Figure 4.**
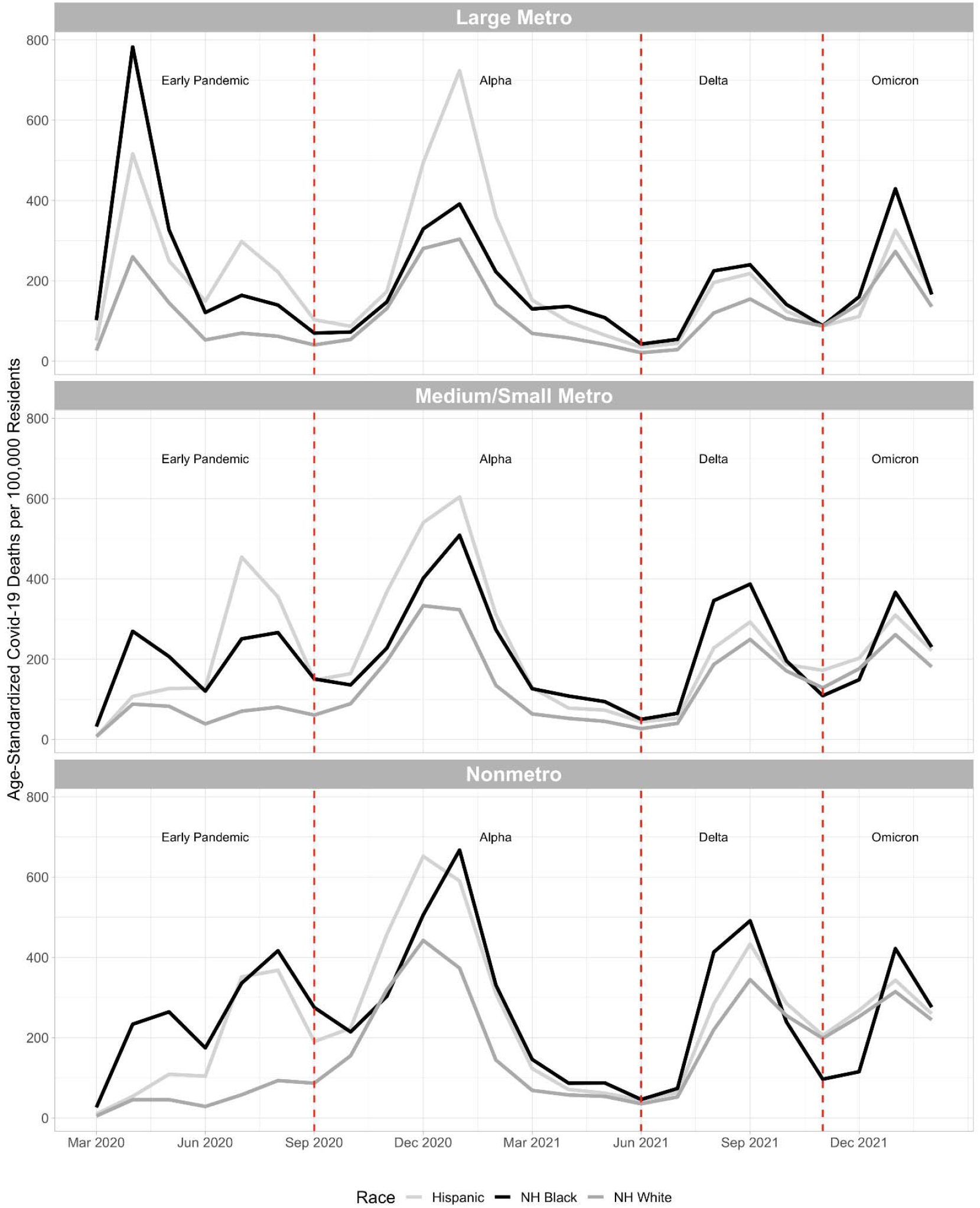
Age-Standardized Covid-19 Death Rates by Race/Ethnicity across Metro-Nonmetro Categories by Month from March 2020 to February 2022 NH = Non-Hispanic <u>Notes:</u> Death rates were age-standardized using 3 age groups (0-54, 55-74 and 75+) and the overall U.S. 2020 population as the standard population. Non-Hispanic NHOPI, Asian, and AIAN were excluded from this figure due to significant data suppression at this level of temporal detail. Data was imputed for one or more age categories for Hispanic residents in nonmetro areas in March 2020 because death counts were between 1 and 9 for one or more age categories and were thus suppressed by CDC WONDER. We divided the pandemic into 4 waves: the early pandemic (March to August 2020), alpha wave (September 2020 to May 2021), delta wave (June to October 2021), and omicron wave (November 2021 to February 2022).

#### Early Pandemic

The early pandemic started with a substantial peak in mortality rates among non-Hispanic Black residents in large metros between March and June 2020. Non-Hispanic Black mortality rates also peaked in medium/small metros and nonmetro areas during this time but at a lower level. Hispanic mortality rates also peaked in large metro areas between March and June 2020 but not to the same levels as non-Hispanic Black mortality. Between June and August 2020, mortality rates peaked among Hispanic residents in metros, medium/small metros, and nonmetro areas, surpassing non-Hispanic Black mortality in metros and medium/small metros. Non-Hispanic white mortality rates remained relatively low and stagnant during the early pandemic, except in large metros where there was a peak between March and June 2020. During the early pandemic, mortality was concentrated in the Northeast and Midwest regions.

#### Alpha Wave

During the alpha wave between October 2020 and March 2021, mortality peaked among Hispanic, non-Hispanic Black, and non-Hispanic white residents across metro-nonmetro categories. In large metros, the peak was greatest for Hispanic residents and smaller for non-Hispanic Black and white residents. In medium/small metros, the peak was greatest for Hispanic residents and smallest for non-Hispanic white residents with non-Hispanic Black residents in between. In nonmetro areas, the peak was equally large among Hispanic and non-Hispanic Black residents and smaller among non-Hispanic white residents. During the alpha wave, the mortality peak was highest in the West.

#### Delta Wave

During the delta wave between June and October 2021, mortality peaked among Hispanic, non-Hispanic Black, and non-Hispanic white residents, with larger peaks occurring in nonmetro areas than medium/small metros and large metros. Across metro-nonmetro categories, non-Hispanic white mortality rates were lower than Hispanic and non-Hispanic Black mortality rates and were higher in nonmetro areas than medium/small metros and large metros. Hispanic and non-Hispanic Black mortality rates followed similar patterns in large metros and nonmetro areas while in medium/small metros, non-Hispanic Black mortality rates were higher than Hispanic mortality rates. During the delta wave, the highest mortality peak was observed in the South.

#### Omicron Wave

During the omicron wave between November 2021 and February 2022, mortality rates peaked among Hispanic, non-Hispanic Black, and non-Hispanic white residents across metro-nonmetro categories. The Non-Hispanic Black mortality rate peaked higher than Hispanic and non-Hispanic white mortality rates in each metro-nonmetro category during this wave. During the omicron wave, mortality was relatively evenly spread across the 4 Census regions, with mortality rates being slightly higher in the Midwest and Northeast.

### Association of Vaccination with Changes in Covid-19 Mortality

Figure 5 examines the relationship between the percent of the overall population fully vaccinated in an area and changes in age-standardized death rates for specific racial/ethnic groups, using combinations of metro-nonmetro categories and Census divisions as the unit of analysis. For Hispanic, non-Hispanic Black, and non-Hispanic white populations, increases in vaccination were associated with reductions in mortality rates during the second year of the pandemic. The reductions were greatest for Hispanic residents (decrease in 4.3 deaths per 100,000 residents per 1 percent increase in the proportion of the population fully vaccinated, R^2^ = 0.21) followed by non-Hispanic white residents (decrease in 3.3 deaths per 100,000 residents per 1 percent increase in the proportion of the population fully vaccinated, R^2^ = 0.45) and non-Hispanic Black residents (decrease in 1.7 deaths per 100,000 residents per 1 percent increase in the proportion of the population fully vaccinated, R^2^ = 0.06). While vaccination was associated with substantial mortality reduction, Hispanic and non-Hispanic Black mortality also appeared to experience a reduction in mortality rates that was not associated with vaccination.

**Figure 5.**
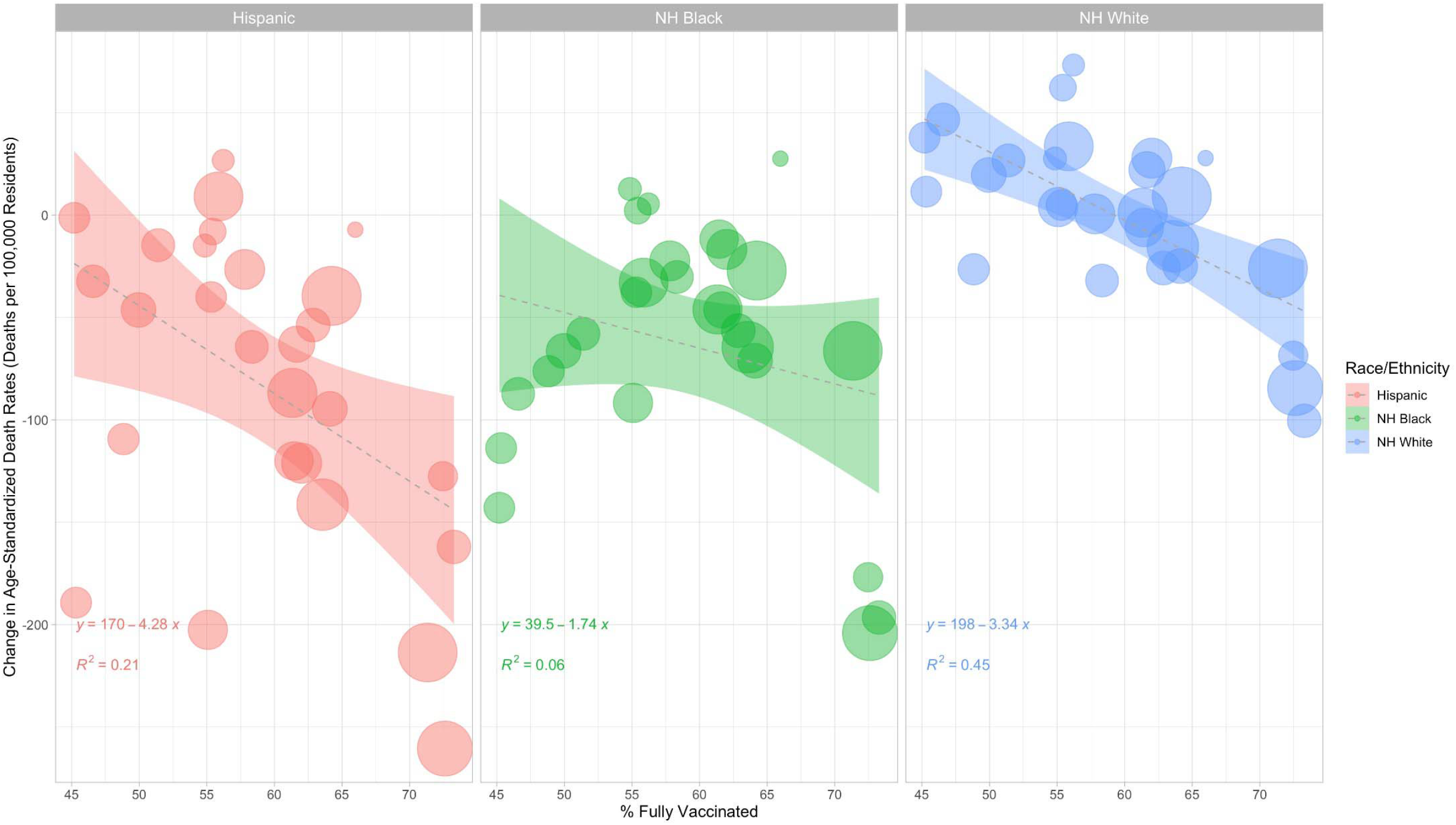
Vaccination and Changes in Age-Standardized Death Rates in Combinations of Metro-Nonmetro Categories and Census Divisions by Race/Ethnicity, March 2020-February 2021 to March 2021-February 2022 NH = Non-Hispanic <u>Notes:</u> Death rates were age-standardized using 3 age groups (0-54, 55-74 and 75+) using the overall U.S. 2020 population as the standard population. The unit in this analysis is combinations of metro-nonmetro categories and Census divisions (i.e. large metro Middle Atlantic). Data was imputed for one or more age categories for Hispanic residents in year 1 and year 2 in nonmetro areas in New England, Hispanic residents in year 1 in nonmetro areas in the Middle Atlantic division, NH Black residents in year 1 in nonmetro areas in the Middle Atlantic division, and NH Black residents in year 1 and year 2 in nonmetro areas in the Mountain division, New England, and the Pacific division because death counts were between 1 and 9 for one or more age categories and were thus suppressed by CDC WONDER. The linear regression lines were weighted using the 2020 population for Hispanic, non-Hispanic Black, and non-Hispanic white residents respectively in each geographic unit.

## DISCUSSION

This study had several key findings. First, when Covid-19 first affected a geographic area, non-Hispanic Black and Hispanic populations experienced extremely high levels of Covid-19 mortality and racial/ethnic inequity that were not repeated at any other time during the pandemic. Second, between the first and second year of the pandemic, racial/ethnic inequities in Covid-19 mortality decreased for Hispanic, non-Hispanic Black, and non-Hispanic AIAN residents across geographic areas due to reductions in mortality rates for these populations and increases in non-Hispanic white mortality rates in some areas. Though racial/ethnic inequities in Covid-19 mortality decreased, substantial inequities still existed in most areas during the second year of the pandemic. Third, non-Hispanic Black, non-Hispanic AIAN, and Hispanic residents reported higher Covid-19 mortality in rural areas than urban areas in the second year of the pandemic, suggesting that these communities are facing distinctive public health challenges. Fourth, non-Hispanic white mortality rates worsened in nonmetro areas during the second year of the pandemic—one of the only instances in the U.S. of Covid-19 mortality increasing in the pandemic’s second year—suggesting there may be some unique factors driving mortality for this population. Finally, vaccination was associated with reductions in Covid-19 mortality between the first and second year of the pandemic, but the effect of population vaccination on mortality may differ by racial/ethnic group.

Prior research has shown that populations with higher social vulnerability and who are more exposed to structural racism are more likely to experience negative health outcomes and mortality during a public health emergency.^49,50^ In this analysis of the Covid-19 pandemic, racial/ethnic inequities in Covid-19 mortality were largest when Covid-19 first spread to a geographic area (i.e. large metros and the Northeast in the early pandemic and nonmetro areas and the West in the alpha wave). This suggests that when public health systems were initially overwhelmed by Covid-19, the greatest effects were on Black residents and other racial/ethnic groups who tend to be negatively impacted by public health policies and systems.

While the Covid-19 pandemic in the U.S. was concentrated heavily in large metros during the early pandemic, it shifted overwhelmingly toward nonmetro areas beginning with the alpha wave. In fact, during the first year of the pandemic, despite the enormous scale of the initial surge in large metro areas, mortality rates were higher in nonmetro areas than large metro areas for non-Hispanic Black, non-Hispanic AIAN, and non-Hispanic white residents and similar between the areas for Hispanic residents. The substantial Covid-19 mortality impact observed in nonmetro areas, especially for Black, Hispanic, and AIAN communities, could suggest that rural health systems were unprepared to respond to the pandemic and that public health measures such as lockdowns and mandatory masking had less of an effect in these communities. While mortality decreased in nonmetro areas for Hispanic, non-Hispanic Black, non-Hispanic AIAN, and non-Hispanic Asian residents during the second year of the pandemic, it remained higher than in medium/small metros and large metros. This rural mortality disadvantage could be explained in part by lower vaccination rates in rural areas. As of January 2022, 61% of metro residents and only 48% of nonmetro residents were fully vaccinated.^51^ Mortality actually increased for non-Hispanic white residents and non-Hispanic NHOPI residents in nonmetro areas and medium/small metros during the second year of the pandemic. In the West and Northeast, mortality also increased among Hispanic, non-Hispanic Black, and non-Hispanic white residents throughout nonmetro areas, suggesting these communities may be areas of public health concern.

Prior research has established that Covid-19 vaccination is an effective public health intervention to reduce Covid-19 mortality.^52,53^ In this study, increased vaccination was associated with reductions in mortality between the first and second year of the pandemic for Hispanic, non-Hispanic Black, and non-Hispanic white populations. The associated reductions were largest among Hispanic residents followed by non-Hispanic white residents and were smallest among non-Hispanic Black residents. This suggests that vaccination may have been more successful in reducing mortality among Hispanic and non-Hispanic white populations than in the non-Hispanic Black population. Differing vaccination rates by race/ethnicity may partially explain this finding. As of July 6, 2022, 54.2% of Hispanic individuals, 49.5% of non-Hispanic white individuals, and 42.7% of non-Hispanic Black individuals have been fully vaccinated.^54^ These disparities are in part due to ineffective vaccine delivery to these communities.^55–57^ We also observed substantial reductions in mortality among non-Hispanic AIAN residents between the first and second year of the pandemic, especially in nonmetro areas. This finding could also be attributed to vaccination rates given that 61.2% of non-Hispanic AIAN individuals have been fully vaccinated, which is higher than other racial/ethnic groups,^54^ and that vaccination campaigns have been particularly successful in nonmetro indigenous communities.^58^ Non-Hispanic Asian residents also have high vaccination rates and reported lower Covid-19 mortality rates than non-Hispanic white residents during the study.^54^

The finding that non-Hispanic white Covid-19 mortality increased substantially in nonmetro areas, medium/small metros, the South, and the West between the first and second year of the pandemic is remarkable. In addition to highlighting the tragic impact of vaccine hesitancy and ineffective vaccine delivery in these communities,^45,51^ this result may also point to the on-going social and structural factors that are influencing contemporary health in this population. Rural populations have higher rates of chronic diseases, increasing their risk of dying from Covid-19,^59^ and rural residents were also likely to engage in protective behaviors such as mask-wearing during the pandemic.^60^ Prior research has also found that automotive plant closures and coal mine closures were associated with increases in mortality among non-Hispanic white residents, demonstrating the negative impact of declining economic opportunity on rural health for white Americans.^61,62^ The Covid-19 pandemic has had significant impacts on economic well-being and employment in rural areas, which may have further reduced rural residents’ access to health care.^63^ Prior research has also established that states with more conservative health policies, such as no Medicaid expansion, low minimum wage, no earned income tax credit, and no paid medical and family leave have shorter life expectancies.^64^ In fact, the U.S. overall has experienced significantly more Covid-19 deaths than other peer countries who have more progressive national and local health policies.^65^ Some of these policies, such as no paid sick leave, may have directly contributed to increased Covid-19 mortality during the pandemic. While the increases in mortality were largest among non-Hispanic white residents in nonmetro areas, other racial/ethnic groups, including non-Hispanic Black, Hispanic, and non-Hispanic AIAN populations, also had elevated mortality rates in nonmetro areas, indicating that most racial/ethnic groups faced substantial health challenges during the pandemic in rural areas and are impacted by the health polices and structural factors affecting rural communities.

While vaccination has been successful in reducing Covid-19 mortality, vaccine delivery in its current form may not fully eliminate racial/ethnic inequities in morality, and additional equity-focused public health policies appear to also be needed.^66^ If vaccination is to reduce and/or eliminate racial/ethnic inequities in mortality, vaccination rates will need to be significantly higher among non-Hispanic Black individuals and other populations with higher Covid-19 mortality rates. Due to medical racism and the legacy of medical experimentation on Black communities and other racial/ethnic groups,^67^ medical mistrust is more common in these communities and has contributed to Covid-19 vaccine hesitancy.^68^ However, adopting equity-promoting vaccination strategies has been shown to increase vaccine uptake in these populations. These approaches include partnerships with faith-based organizations, housing communities, and trusted organizations led by community members; mitigating barriers such as removing requirements for photo identification; making vaccines available outside typical working hours; ensuring that clinics are accessible via public transportation; and centering public health officials of color, in particular Black officials, in the design and implementation of outreach efforts and publicly presenting these officials at the forefront of national vaccine communication.^47,67,69–71^ These strategies may be applied to both primary series vaccination and subsequent boosters. It is particularly important that these strategies are adapted to both urban and rural contexts, given the rural mortality disadvantage observed in this study. Vaccination efforts early in the pandemic also prioritized older populations, which favored non-Hispanic white residents since the age distribution of the non-Hispanic white population is older than other racial/ethnic groups.^72,73^ Instead, a vaccination strategy that targets geographic areas with high Covid-19 death rates and populations with more comorbidities could save more lives.^74^

In addition to equitable vaccine delivery, additional public health policies are urgently needed to intervene on each of the mechanisms through which structural racism is operating during the pandemic: infection, weathering, and experiences in health care. First, differential rates of Covid-19 infection can be modified by addressing occupational segregation and household crowding. Paid sick leave and paid medical and family leave would ensure that essential workers are able to appropriately isolate if they develop Covid-19 symptoms and have adequate time to recover if they test positive and become seriously ill.^32,75^ Furthermore, workplace safety precautions such as the provision of effective masks and monitoring of workplace air circulation are needed. Regarding household crowding, rent/eviction moratoriums, mortgage relief moratoriums and extended Covid-19 unemployment benefits would prevent crowding related to economic and housing insecurity.^76,77^ Second, although underlying health risks for Covid-19 resulting from weathering may not be possible to fully reverse, future harm can be prevented. Policies that would advance racial health equity, such as economic reparations, divestments from police funding and investments in community-based safety programs, and increased funding for community and social programs in underserved communities, are needed.^30,78,79^ Finally, racial disparities in health insurance could be addressed through Medicaid Expansion at the state-level or a national universal health insurance option that would address the remaining costs of co-pays, prescriptions, premiums, and high deductibles.^80,81^ Investments in diversifying the physician workforce, increased training in the provision of culturally competent care, and policies to hold medical providers responsible for medical racism would also make a long-term impact on the readiness of the public health system to equitably respond to a pandemic.^82,83^

### Limitations

This study had several limitations. First, NCHS mortality data for 2021 used in this study was provisional and was thus incomplete and subject to revision. Second, population estimates for 2022 were not available at the time of our analysis, and some geographic units may have experienced population shifts which we could not account for. Third, we used a data imputation procedure (for less than 5% of data) to replace data that the CDC suppressed because the count was between 1 and 9 deaths. Fourth, we used 3 age categories rather than 10-year age categories for age-standardization due to data suppression, especially among the less populous racial/ethnic groups. Fifth, we did not assess the multi-racial category because we assumed it would not represent a homogeneous population across the United States. Future research can avoid this challenge by using bridged race categories once they become available for 2021 and 2022 mortality data. Finally, this study represents an analysis of Covid-19 mortality rather than excess mortality, which would capture undiagnosed Covid-19 deaths and deaths indirectly related to the pandemic. Since excess deaths not assigned to Covid-19 may have occurred differentially by race/ethnicity, the mortality patterns observed in this paper may look different when examined using excess mortality. For example, prior research has shown that excess deaths not assigned to Covid-19 are more common in Black communities.^3^ Future studies could repeat this analysis of geographic patterns using measures of excess mortality.

## CONCLUSION

This study builds on the substantial prior literature on racial/ethnic inequities in Covid-19 mortality by identifying geographic areas where mortality rates have increased and decreased between the first and second years of the pandemic for various racial/ethnic groups. Vaccination rates were associated with reductions in Covid-19 morality for Hispanic, non-Hispanic Black, and non-Hispanic white residents, and increased vaccination may have contributed to the decreases in racial/ethnic inequities in Covid-19 mortality observed in the second year of the pandemic. Despite these reductions, substantial inequities remain that require urgent health policy action. Mortality is elevated in nonmetro areas and increasing for some racial/ethnic groups, highlighting the need for increased vaccine delivery and other public health measures in rural communities. Taken together, these findings highlight the continued need to prioritize health equity in the pandemic response and to modify the structures and policies through which systemic racism operates and has generated racial health inequities.

## Data Availability

Data used in the present manuscript are publicly available from the Centers for Disease Control and Prevention and U.S. Census Bureau.

## SUPPLEMENTAL TABLES/FIGURES

**Table S1.**
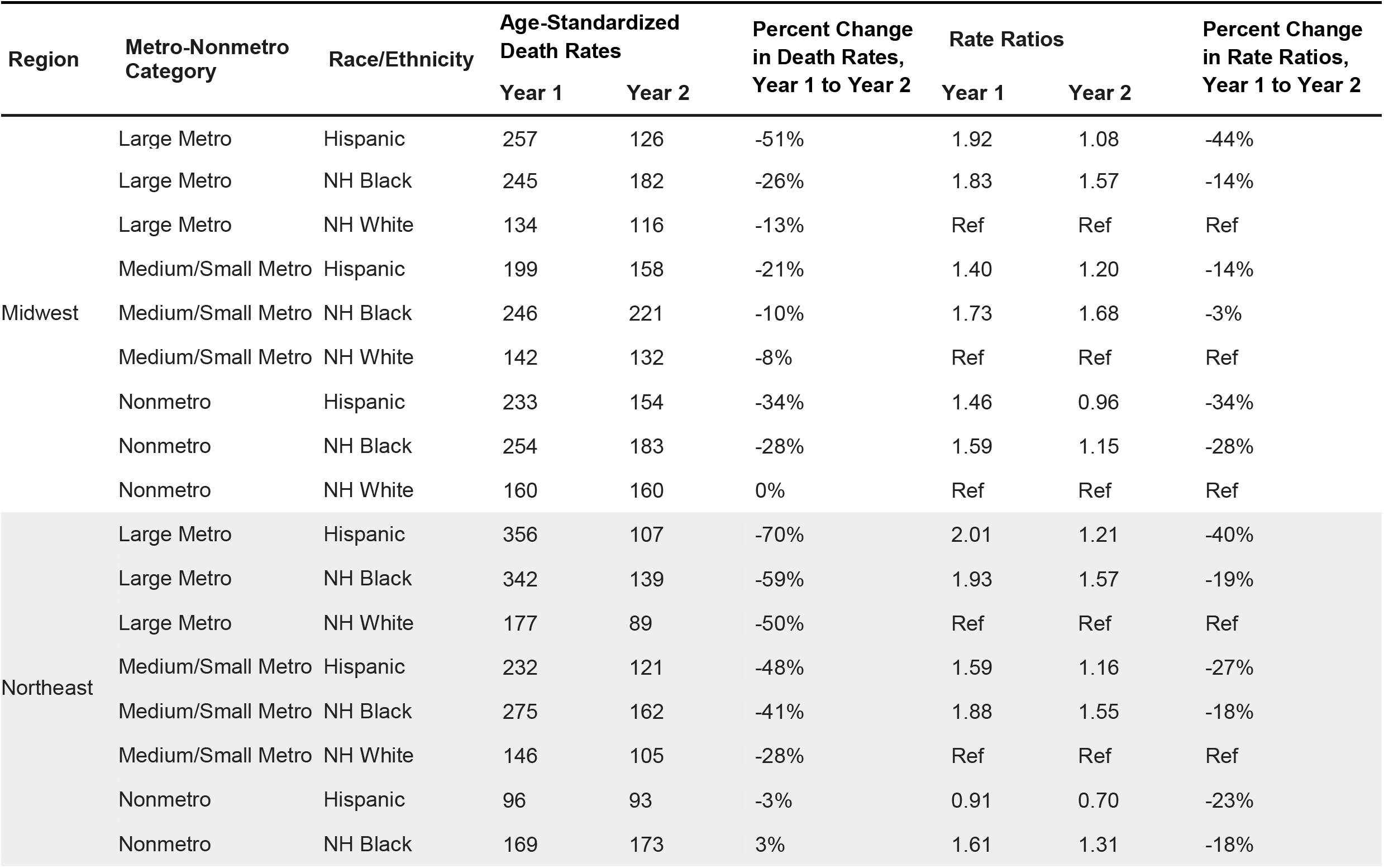
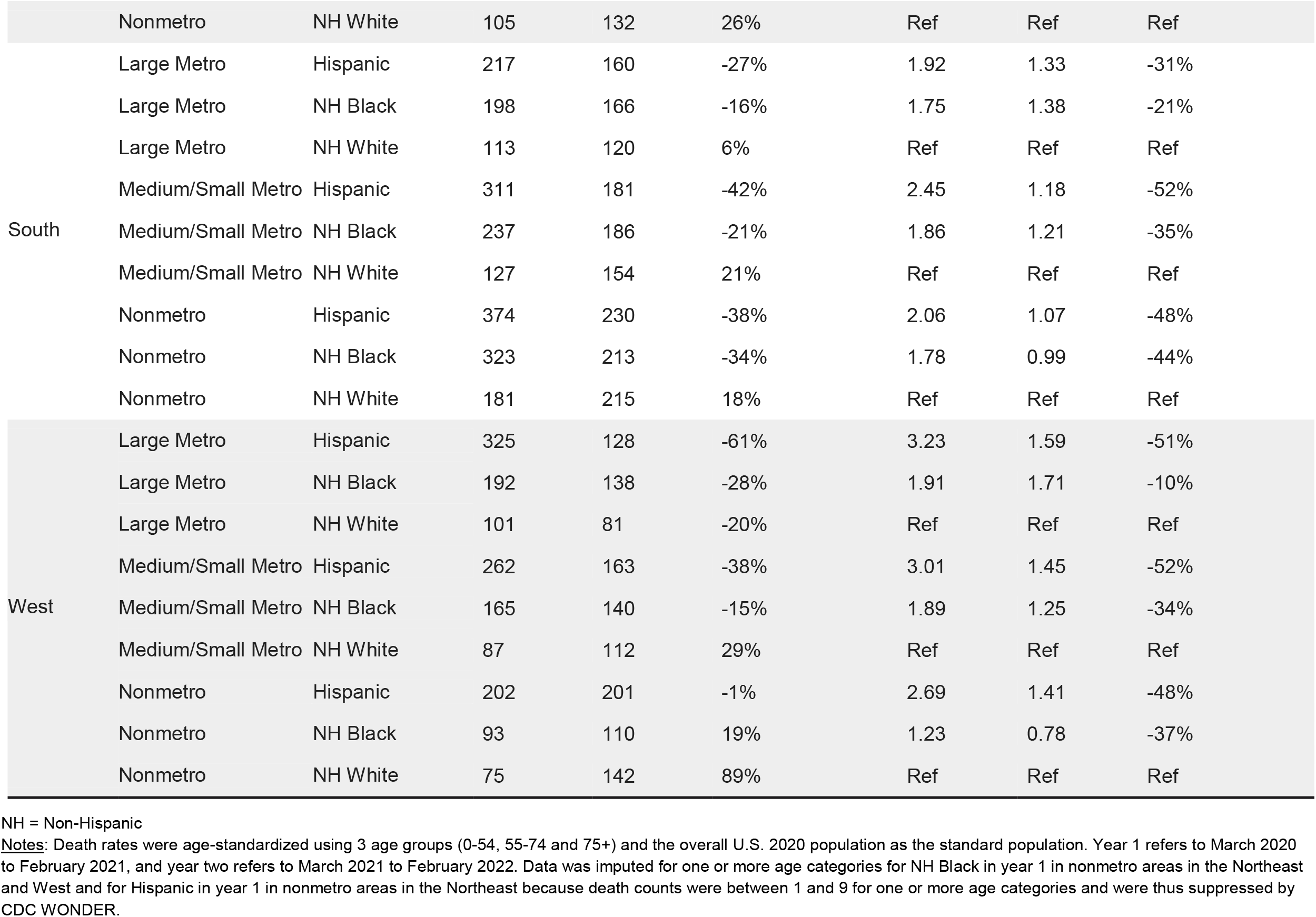
Changes in Age-Standardized Covid-19 Death Rates by Race/Ethnicity across Combinations of Census Regions and Metro-Nonmetro Categories, March 2020-February 2021 to March 2021-February 2022

**Figure S1.**
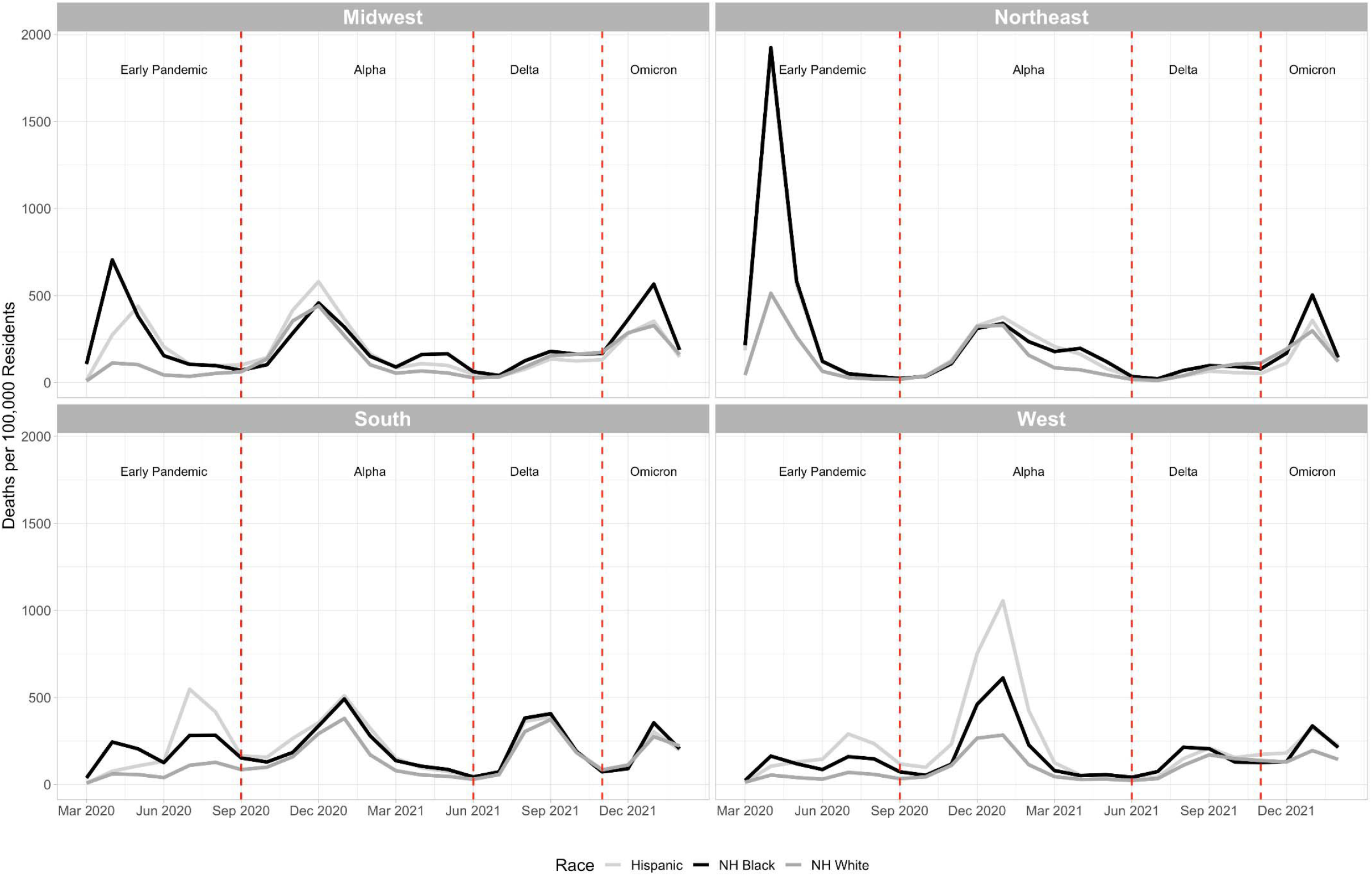
Age-Standardized Covid-19 Death Rates by Race/Ethnicity across Census Regions by Month from March 2020 to February 2022 NH = Non-Hispanic <u>Notes:</u> Death rates were age-standardized using 3 age groups (0-54, 55-74 and 75+) and the overall U.S. 2020 population as the standard population. Data was imputed for one or more age categories for Hispanic residents in the Midwest in March 2020 because death counts were between 1 and 9 for one or more age categories and were thus suppressed by CDC WONDER.

## METHODS SUPPLEMENT

### Data Sources & Extraction Procedure

#### Mortality Data

##### Data Source

CDC WONDER (https://wonder.cdc.gov/mcd-icd10-provisional.html)

##### Rationale for Querying Approach

While it is possible to query CDC WONDER for mortality data by sorting with variables such as ethnicity, race, 10-year age groups, urbanization, region, month, and year, sorting by these variables reduces the data into smaller cells than required for the analysis, resulting in unnecessary data suppression since death counts between 1 and 9 are suppressed. Thus, it is critical to submit separate queries for each piece of data needed so that no unnecessary suppression occurs, especially for racial/ethnic groups with smaller populations. In total, 192 CDC WONDER queries are needed to extract all the data needed for this analysis.

##### CDC WONDER Query #1 (Table 1 and Figure 1)

Covid-19 Mortality by Race/Ethnicity, Urbanization and Year (36 Files Total)

- Use Multiple Cause Provisional Mortality Statistics, 2018 through Last Month
- Select Multiple Cause of Death (Any Mention on Death Certificate): Covid-19 (U07.1)
- Select Show Zero Values and Suppressed Values
- Select Months (Separate Files for Each)
  - March 2020 to February 2021
  - March 2021 to February 2022
- Select 2013 Urbanization (by Residence) Categories (Separate Files for Each)
  - Nonmetro (Noncore (Nonmetro) and Micropolitan (Nonmetro))
  - Small/Medium Metro (Small Metro and Medium Metro)
  - Large Metro (Large Fringe Metro and Large Central Metro)
- Select 10-Year Age Categories (Separate Files for Each)
  - Ages 0-54
  - Ages 55-74
  - Ages 75+
- Select Race/Ethnicity (Separate files for Each)
  - Select Hispanic and All Races
  - Select Non-Hispanic and All Races and Group by Single Race 6

##### CDC WONDER Query #2 (Table 1 and Figure 1)

Covid-19 Mortality by Race/Ethnicity, Census Region and Year (12 Files Total)

- Use Multiple Cause Provisional Mortality Statistics, 2018 through Last Month
- Select Multiple Cause of Death (Any Mention on Death Certificate): Covid-19 (U07.1)
- Select Show Zero Values and Suppressed Values
- Select Months (Separate Files for Each)
  - March 2020 to February 2021
  - March 2021 to February 2022
- Select 10-Year Age Categories (Separate Files for Each)
  - Ages 0-54
  - Ages 55-74
  - Ages 75+
- Specify Race/Ethnicity (Separate files for Each)
  - Select Hispanic and All Races
  - Select Non-Hispanic and All Races and Group by Single Race 6
- Group by Census Region (by Residence)

##### CDC WONDER Query #3 (Figure 2 and Table S1)

Covid-19 Mortality by Race/Ethnicity, Census Region, Metro-Nonmetro Category and Year (54 Files Total)

- Use Multiple Cause Provisional Mortality Statistics, 2018 through Last Month
- Select Multiple Cause of Death (Any Mention on Death Certificate): Covid-19 (U07.1)
- Select Show Zero Values and Suppressed Values
- Select Months (Separate Files for Each)
  - March 2020 to February 2021
  - March 2021 to February 2022
- Select 2013 Urbanization (by Residence) Categories (Separate Files for Each)
  - Nonmetro (Noncore (Nonmetro) and Micropolitan (Nonmetro))
  - Small/Medium Metro (Small Metro and Medium Metro)
  - Large Metro (Large Fringe Metro and Large Central Metro)
- Select 10-Year Age Categories (Separate Files for Each)
  - Ages 0-54
  - Ages 55-74
  - Ages 75+
- Specify Race/Ethnicity (Separate files for Each)
  - Select Hispanic and All Races
  - Select Non-Hispanic and Black or African American
  - Select Non-Hispanic and White
- Group by Census Region

##### CDC WONDER Query #4 (Figure 4)

Covid-19 Mortality by Race/Ethnicity, Urbanization and Month (27 Files Total)

- Use Multiple Cause Provisional Mortality Statistics, 2018 through Last Month
- Select Multiple Cause of Death (Any Mention on Death Certificate): Covid-19 (U07.1)
- Select Show Zero Values and Suppressed Values
- Select Months March 2020 to February 2022
- Select 2013 Urbanization (By Residence) Categories (Separate Files for Each)
  - Nonmetro (Noncore (Nonmetro) and Micropolitan (Nonmetro))
  - Small/Medium Metro (Small Metro and Medium Metro)
  - Large Metro (Large Fringe Metro and Large Central Metro)
- Select 10-Year Age Categories (Separate Files for Each)
  - Ages 0-54
  - Ages 55-74
  - Ages 75+
- Select Race/Ethnicity (Separate files for Each)
  - Select Hispanic and All Races
  - Select Non-Hispanic and Black or African American
  - Select Non-Hispanic and White
- Group by Month

##### CDC WONDER Query #5 (Figure 3 and Figure 5)

Covid-19 Mortality by Race/Ethnicity, Census Division, Metro-Nonmetro Category and Year (54 Files Total)

- Use Multiple Cause Provisional Mortality Statistics, 2018 through Last Month
- Select Multiple Cause of Death (Any Mention on Death Certificate): Covid-19 (U07.1)
- Select Show Zero Values and Suppressed Values
- Select Months (Separate Files for Each)
  - March 2020 to February 2021
  - March 2021 to February 2022
- Select 2013 Urbanization (By Residence) Categories (Separate Files for Each)
  - Nonmetro (Noncore (Nonmetro) and Micropolitan (Nonmetro))
  - Small/Medium Metro (Small Metro and Medium Metro)
  - Large Metro (Large Fringe Metro and Large Central Metro)
- Select 10-Year Age Categories (Separate Files for Each)
  - Ages 0-54
  - Ages 55-74
  - Ages 75+
- Specify Race/Ethnicity (Separate files for Each)
  - Select Hispanic and All Races
  - Select Non-Hispanic and Black or African American
  - Select Non-Hispanic and White
- Group by Census Division (By Residence)

##### CDC WONDER Query #6 (Figure S1)

Covid-19 Mortality by Race/Ethnicity, Region and Month (9 Files Total)

- Use Multiple Cause Provisional Mortality Statistics, 2018 through Last Month
- Select Multiple Cause of Death (Any Mention on Death Certificate): Covid-19 (U07.1)
- Select Show Zero Values and Suppressed Values
- Select Months March 2020 to February 2022
- Select 10-Year Age Categories (Separate Files for Each)
  - Ages 0-54
  - Ages 55-74
  - Ages 75+
- Select Race/Ethnicity (Separate files for Each)
  - Select Hispanic and All Races
  - Select Non-Hispanic and Black or African American
  - Select Non-Hispanic and White
- Group by Census Region (By Residence) and Month

#### Population Data

##### Data Source

U.S. Census 2021 Vintage Population Estimates (https://www2.census.gov/programs-surveys/popest/datasets/2020-2021/counties/asrh/cc-est2021-all.csv)

Data Dictionary (https://www2.census.gov/programs-surveys/popest/technical-documentation/file-layouts/2020-2021/cc-est2021-alldata.pdf)

This file provides county-level estimates of population by race/ethnicity and age, which can be aggregated to the levels of metro-nonmetro categories, Census regions, and states. Querying CDC WONDER by county and metro-nonmetro category, Census region, or state will generate the necessary crosswalks for aggregation.

Non-bridged race categories (Hispanic, non-Hispanic AIAN alone, non-Hispanic Asian alone, non-Hispanic NHOPI alone, non-Hispanic Black alone, and non-Hispanic white alone) were used for this analysis. Bridged race estimates are not yet available for 2021 and 2022 mortality data.

#### Vaccination Data

##### Data Source

CDC Covid-19 Vaccination Data by County (https://data.cdc.gov/Vaccinations/COVID-19-Vaccinations-in-the-United-States-County/8xkx-amqh/data)

The relevant variable is percent of the county fully vaccinated (Series_Complete_Pop_Pct), as of February 28, 2022.

### Age-Standardization Procedure

The U.S. 2020 population was used as the standard population. 70.3% of the population was between the ages of 0 to 54 years, 22.7% was between the ages of 55 to 74 years, and 7.0% was 75 years or older.

For each geographic and temporal unit, age-specific death rates were calculated for ages 0-54, 55-74, and 75+. Each age-specific death rate was multiplied by the corresponding percentage of the population in that age group, and then the 3 components were added together to produce the age-standardized death rate.

It was not possible to use 10-year age intervals for age-standardization due to data suppression.

